# The Impact of the Selective Cytopheretic Device on Neutrophil-to-Lymphocyte Ratios and Hematological Parameters in AKI: A Pooled Analysis

**DOI:** 10.1101/2025.01.14.25320534

**Authors:** Sai Prasad N. Iyer, Nicholas J. Ollberding, Jay L. Koyner, Lenar T. Yessayan, Kevin K. Chung, H. David Humes

## Abstract

**Background:** The Selective Cytopheretic Device (SCD) is an immunomodulatory cell-directed extracorporeal therapy that reprograms activated neutrophils and monocytes towards immune homeostasis in hyperinflammatory conditions such as acute kidney injury (AKI). However clinical mechanisms remain unclear.

**Methods:** We examined the effect of SCD treatment from prior AKI clinical studies on systemic inflammation through neutrophil-to-lymphocyte ratios (NLR) and other hematological measures to gain insights into the mechanism of the SCD. Linear-mixed effects regression was used to estimate differences in NLR and other hematological measures between SCD treated patients and controls over the first six days after initiating CKRT.

**Results:** Hematological data were analyzed from 98 patients with AKI requiring continuous kidney replacement therapy (CKRT) treated with the SCD, and 32 CKRT only control patients. SCD reduced NLR across all individual studies through Day 6 of treatment, while the control group demonstrated upward trends in NLR past day 3. When analyzed as pooled groups, both cohorts displayed similar baseline NLRs (SCD = 23.6 vs. control = 21.7; p=0.636). SCD treated adult patients demonstrated a statistically significant reduction in NLR vs. control adult patients at Day 6 (SCD = 13.3 vs. control = 25.7 at day 6; p_trend_ = 0.011). This difference was maintained following sensitivity analysis upon exclusion of an adult ICU study due to a shorter follow-up period (SCD = 13.7 vs. control = 25.6; p_trend_ = 0.013). NLR reductions in the SCD group were driven by decreases in neutrophils and increases in lymphocytes. No statistically significant differences were observed between groups for monocyte-to-lymphocyte ratio (MLR) levels or platelets over the same treatment duration.

**Conclusions:** In a pooled analysis of multiple AKI clinical studies SCD treatment demonstrated reductions in NLR. This analysis provides further clinical mechanistic evidence of leukocyte immunomodulation in targeting the dysregulation of effector immune cells in hyperinflammatory conditions such as AKI and sepsis.

## Introduction

The selective cytopheretic device (SCD) represents a novel class of immunomodulatory cell-directed extracorporeal therapies that is integrated with continuous kidney replacement therapy (CKRT) under regional citrate anticoagulation (RCA) in the treatment of AKI in both adult and pediatric patients^1–3^. Recently it has been approved by FDA under a humanitarian device exemption (HDE) to treat children with AKI due to sepsis or a septic condition^4,5^. The SCD’s unique membrane microenvironment, characterized by a low shear stress state mimicking physiologic conditions seen in capillary bed, and a low ionized calcium (iCa) environment (<0.4 mM) facilitated by RCA, enables it to selectively target the most highly activated neutrophils and monocytes. After binding to SCD membranes, pro-inflammatory, activated neutrophils are deactivated and reprogrammed for apoptosis, while proinflammatory monocytes are shifted toward a less inflammatory, reparative phenotype within the device. The selectively reprogrammed cells are then released back into the systemic circulation (distinguishing it from adsorptive columns or depletive filters), in the hopes of promoting the natural reparative process towards immune homeostasis and recovery^2,3,6,7^. The SCD achieves immunomodulation without causing immunosuppression or increasing the risk of infections^8^. However, the exact mechanism by which the SCD shifts an imbalanced immune response toward homeostasis is incompletely understood^6^.

Relationships such as neutrophil-to-lymphocyte ratios (NLR) have emerged over the last decade as being a more insightful indicator of systemic inflammation than conventional measures such as CRP since it provides information on components of both innate and adaptive immunity. NLR has prognostic value and independently correlates with mortality and disease progression in the general population and across many diseases (e.g., sepsis, AKI, COVID-19, cancer, cardiovascular and neurodegenerative diseases, inflammatory bowel disease etc.)^9–27^. Specifically, within the AKI and sepsis, and for patients on CKRT, several studies have demonstrated significant positive correlations between a high NLR and worsening disease severity, progression, and clinical outcomes^9,25–27^. The SCD has demonstrated clinical effectiveness with improved outcomes across multiple adult and pediatric AKI studies^7,28–33^. To further understand the immunomodulatory mechanism of the SCD, we analyzed several hematological measures and immune cell relationships, including NLR and monocyte-to-lymphocyte ratio (MLR) and their responses to treatment over time in SCD-treated patients compared to controls from previous AKI studies.

## Methods

Complete blood count with differential (CBC diff), NLR, MLR, and platelets were included from available clinical data from six prior studies in patients with AKI requiring continuous kidney replacement therapy (CKRT). These studies comprised of adult (China Pilot study, ARF-002, SCD-003, SCD-005)^7,28–30^ and two pooled pediatric studies (SCD-PED)^31–33^. **Supplementary Table 1** provides a listing of the studies included, breakdown of patients, and their clinical outcomes. CBC differential data was available for a total of 98 patients who received the SCD across the 4 adult and 2 pediatric studies. CBC differential data for the sensitivity analysis of all pooled adult studies (excluding the pediatric studies) was available for n = 76 patients. The control CKRT only group included patients from a randomized control trial (SCD-003 riCa control group, n=28)^29^, and a contemporaneous matched cohort (CRRTnet; n=4)^34^, resulting in a total sample size of n = 32. See **Figure 1** for the CONSORT flow diagram of included patients. Patients were treated for up to 10 days with SCD + CKRT using RCA. NLRs were calculated by dividing the absolute neutrophil count (ANC) by the absolute lymphocyte counts (ALC). Monocyte-to-lymphocyte ratios (MLR) were calculated by dividing the absolute monocyte count by ALC in the pooled adult studies; however monocyte data from study SCD-005 were not available for inclusion in the analysis. Similarly, platelet counts were also analyzed across the pooled adult studies (except for SCD-005 due to the abovementioned lack of available data). Linear mixed effects regression (LMER) was used to estimate differences in NLR and other hematological measures for SCD-treated patients compared to controls over the first six days after initiating CKRT. Six days post therapy was chosen to ensure that estimates at later time points were not driven by the subset of studies with longer duration given the variability in follow-up across studies.

**Figure 1:**
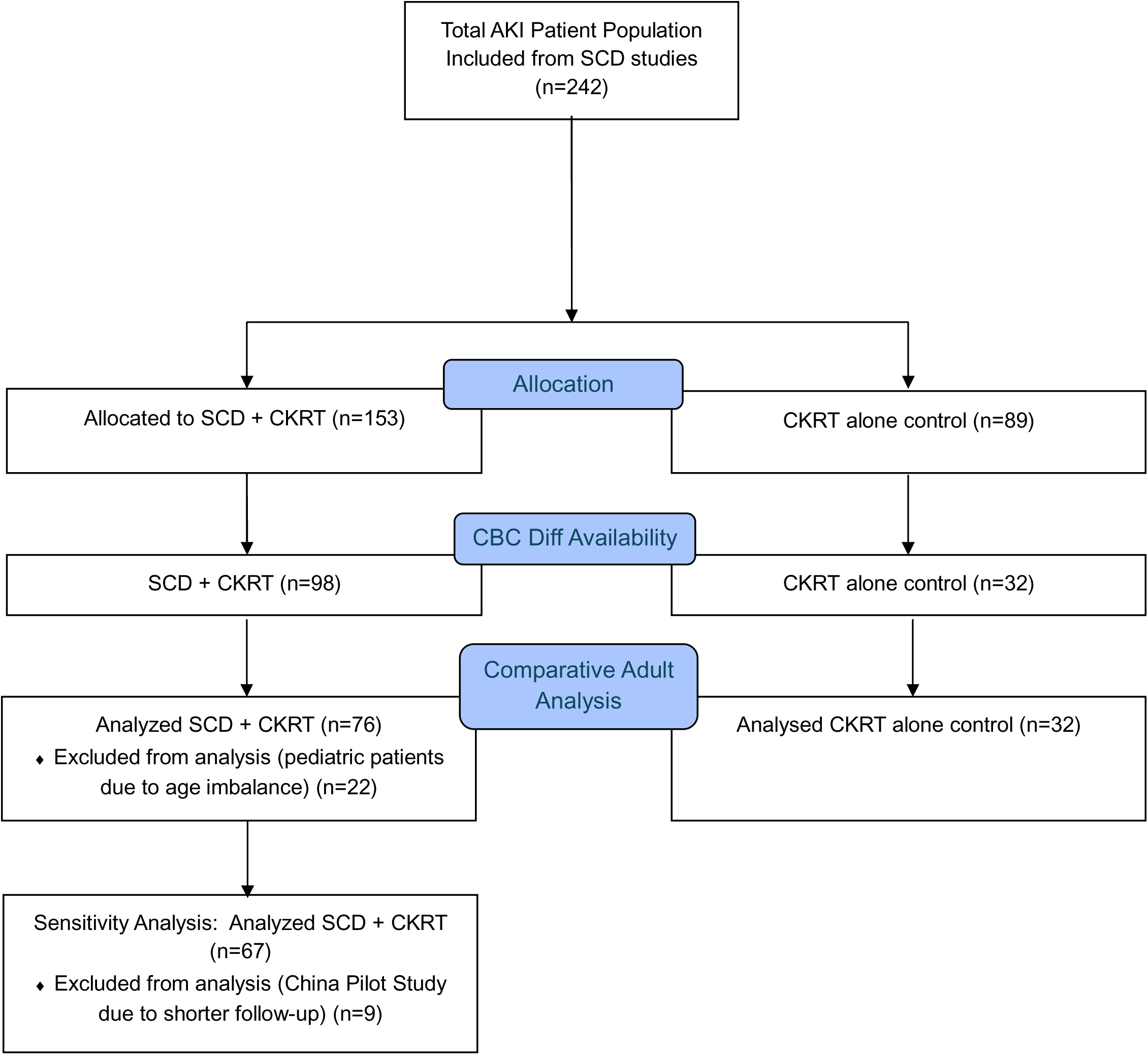
**Consort Flow Diagram of Patients analyzed for Hematological Analysis**

The models included a Gaussian link function to model the continuous responses; random subject-specific intercepts and slopes to allow the values at baseline and the trends over time to differ for each subject; and fixed effect terms for time, treatment group, and the time-by-treatment interaction. Time was modeled using restricted cubic spline terms with knots placed at the 10^th^, 50^th^, and 90^th^ percentiles, using the default settings in the rms (version 6.7.0)^35^ package in R. Likelihood ratio tests of nested models were used to obtain p-values for the differences in response profiles between SCD and control patients. Marginal effect plots were used to visualize the NLR values over time. All analyses were conducted using the R software environment for statistical computing and graphics version (4.1.1)^36^. LMER models were fitted using the glmmTMB package (version 1.1.2.3)^37^.

## Results

A total of 153 patients treated with the SCD were included in the analysis plan across the 6 prior adult and pediatric AKI studies with the SCD. Four of these six studies were single-arm studies. Control patients were obtained from two adult studies (SCD-003 and SCD-005), resulting in a total of 89 control patients for consideration. **Supplementary Tables 1** and **2** summarize the studies, their main clinical outcomes, and the individual patient baseline characteristics. To address the imbalance in patient numbers between the control and SCD cohorts, we aggregated data from all adult studies, excluding pediatric cohorts due to age discrepancies. We then compared the available clinical characteristics between the SCD and control patients in the remaining studies to ensure that the baseline distribution of demographic and clinical characteristics was similar between the two groups (**Table 1)**. No material differences in baseline clinical characteristics were observed between the adult SCD and control cohorts, supporting intergroup comparisons. Patients with AKI requiring CKRT across the pooled adult studies were critically ill, with a majority having sepsis (74.8% in the SCD group vs. 71.8% in the control group; p=0.36), multi-organ failure with high SOFA scores (12.6 in the SCD group vs. 13.1 in the control group; p=0.29), and requiring mechanical ventilation (90.1% in the SCD group vs. 88.2% in the control group; p=0.21) in both the SCD and control groups. Most patients (>60%) in the adult AKI studies were on vasoactive medications (**Supplementary Table 2**). Pediatric patients with AKI from the SCD-PED studies were of similar disease severity, with 68% having sepsis, 95% requiring MV and nearly 64% on vasoactive medications.

**Table 1:** Pooled Adult Baseline Mean Patient Demographics.

| # of Patients per Study | SCD (N=131) | Control (N=89) | P-value |
| --- | --- | --- | --- |
| Age (years), mean (SD) | 56.4 (14.4) | 54 (14.5) | 0.23 |
| Female, n (%) | 47 (35.9) | 33 (38.8) | 0.39 |
| Race, n (%) |  |  | 0.09 |
| Asian | 9 (6.9) | 0 (0) |  |
| Black | 26 (19.8) | 19 (22.4) |  |
| White | 95 (72.5) | 59 (69.4) |  |
| Other | 5 (3.8) | 3 (3.5) |  |
| COVID-19, n (%) | 22 (16.8) | 16 (18.8) | 0.58 |
| SOFA score, mean (SD) | 12.6 (3.3) | 13.1 (3.5) | 0.29 |
| MV, n (%) | 118 (90.1) | 75 (88.2) | 0.21 |
| ECMO, n (%) | 9 (6.9) | 4 (4.7) | 0.57 |
| Sepsis, n (%) | 98 (74.8) | 61 (71.8) | 0.36 |

During our review of available clinical data for hematological analysis, we identified 98 SCD patients and 32 control patients with CBC diff samples with available ANC, ALC, AMC and platelet levels. **Figure 2** illustrates the effect of SCD treatment on NLR over time across each adult and the pooled pediatric studies. Baseline NLRs were mean ± SD: 22.4 ± 20.9 in the adult studies and lower in the pooled pediatric studies 10.9±6.9. **Figure 2A** shows that SCD treatment consistently resulted in downward slopes of NLR across all studies during the first 6 days of therapy in the SCD group except for SCD-005. In contrast, the trend in the control group in SCD-003 shows an initial downward slope which then shifts upward, indicating an increase in NLR as time progresses on CKRT alone (**Figure 2B)**. Control patients in SCD-005 displayed an upward trend in NLR slope throughout their entire course of therapy.

**Figure 2:**
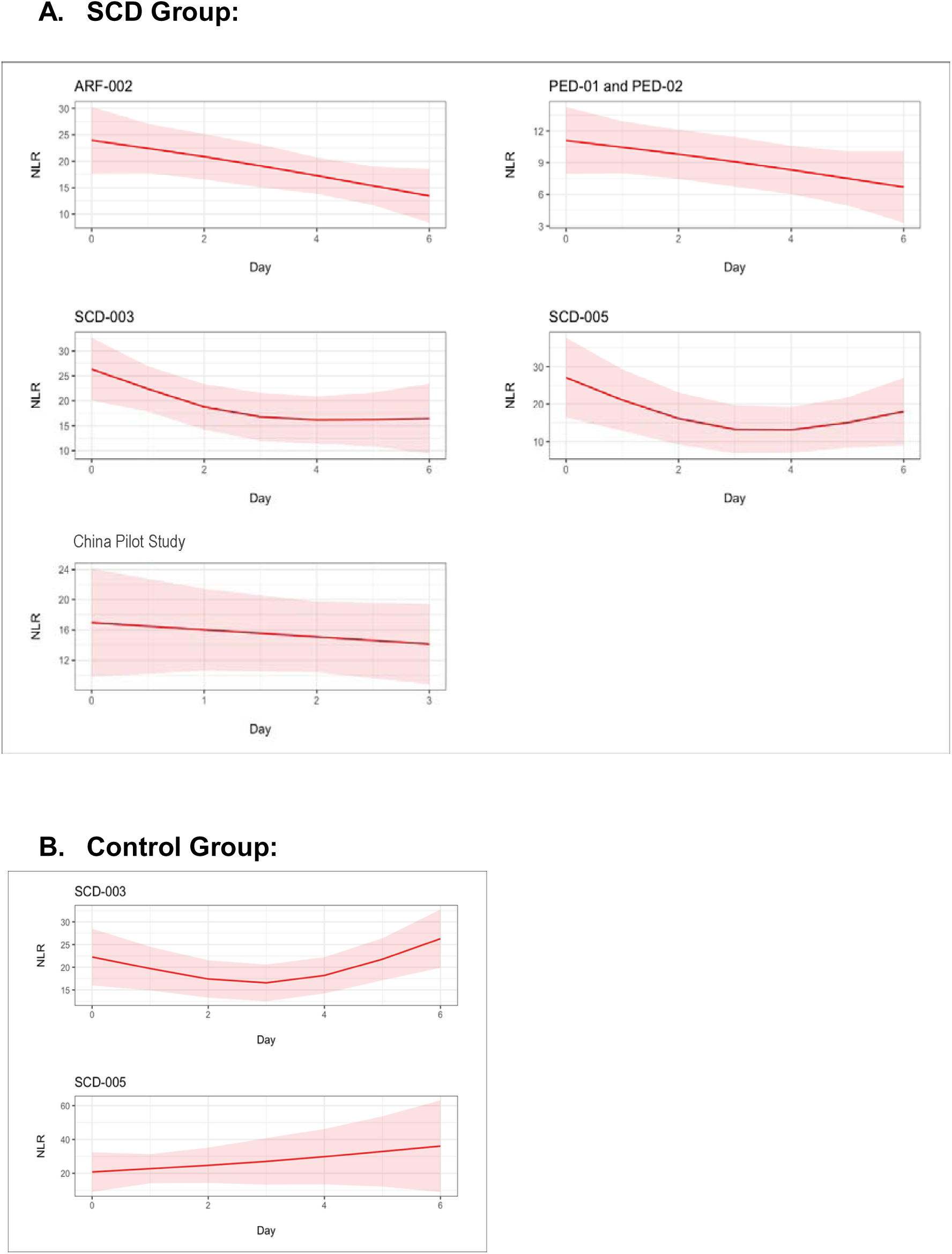
**Linear mixed effect model of the effect of SCD on NLR over time in (A) SCD treated vs. (B) Control patients across individual adult and pediatric AKI studies.**

Next, we examined differences in the observed trends in NLR between the SCD and control groups. To increase statistical power and the precision of our estimates, we combined the SCD and control groups across all adult AKI studies (excluding the pediatric studies due to age imbalances). Exclusion of pediatric patients resulted in a final pooled adult patient population of 76 patients, yielding a ratio of just over 2:1 of SCD: control group (n=32) for comparison (**Figure 1**). **Figure 3A** shows the comparison in trends between the two groups. Both groups exhibited similar NLRs at the beginning of treatment on Day 0, indicating comparable baseline levels of systemic inflammation (SCD = 23.6, control = 21.7, p = 0.636). However, following treatment with SCD, a distinct divergence in NLR trends emerged between the groups. By or shortly after Day 2, the SCD group showed a sustained decline in NLR until Day 6, whereas the control group experienced a reversal in trend shortly after Day 3, with an increase in NLR by Day 6 (SCD = 13.3 vs. control = 25.7, p=0.002; p_trend_=0.011). This represented a net difference of −10.3 in the SCD group compared to +4 in the control group for NLR point estimates from Day 0 to Day 6. Sensitivity analysis excluding the China Pilot study showed that the interaction remained largely unchanged (SCD = 13.7 vs. control = 25.6, p = 0.002 at day 6; p_trend_=0.013; **Figure 3B**), confirming the consistency of the observation that SCD treatment reduces NLR in patients with AKI. NLR declines in the pooled adult population were due to steady decreases in ANC through Day 6, along with continued increases in ALC in the SCD group (**Figure 4**, left top and bottom panels respectively). Upward NLR trends in the control group, however, were due to a mostly static ANC, coupled with initial increases in ALC until about day 3, after which a downward shift in the curve was observed (**Figure 4**, right top and bottom panels respectively).

**Figure 3A:**
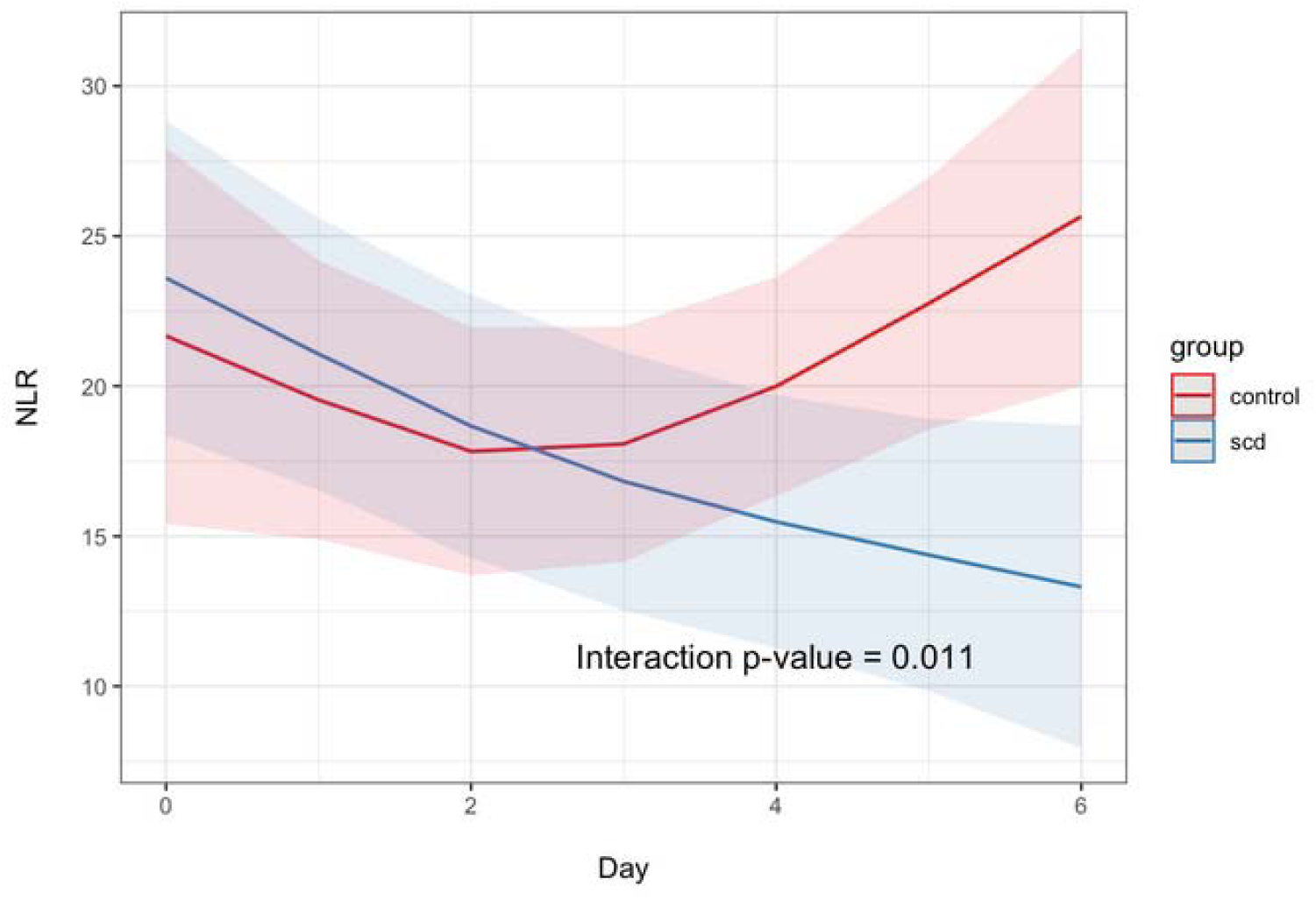
**Effect of SCD treatment on NLR in pooled SCD vs. control patients across the adult AKI studies.**

**Figure 3B:**
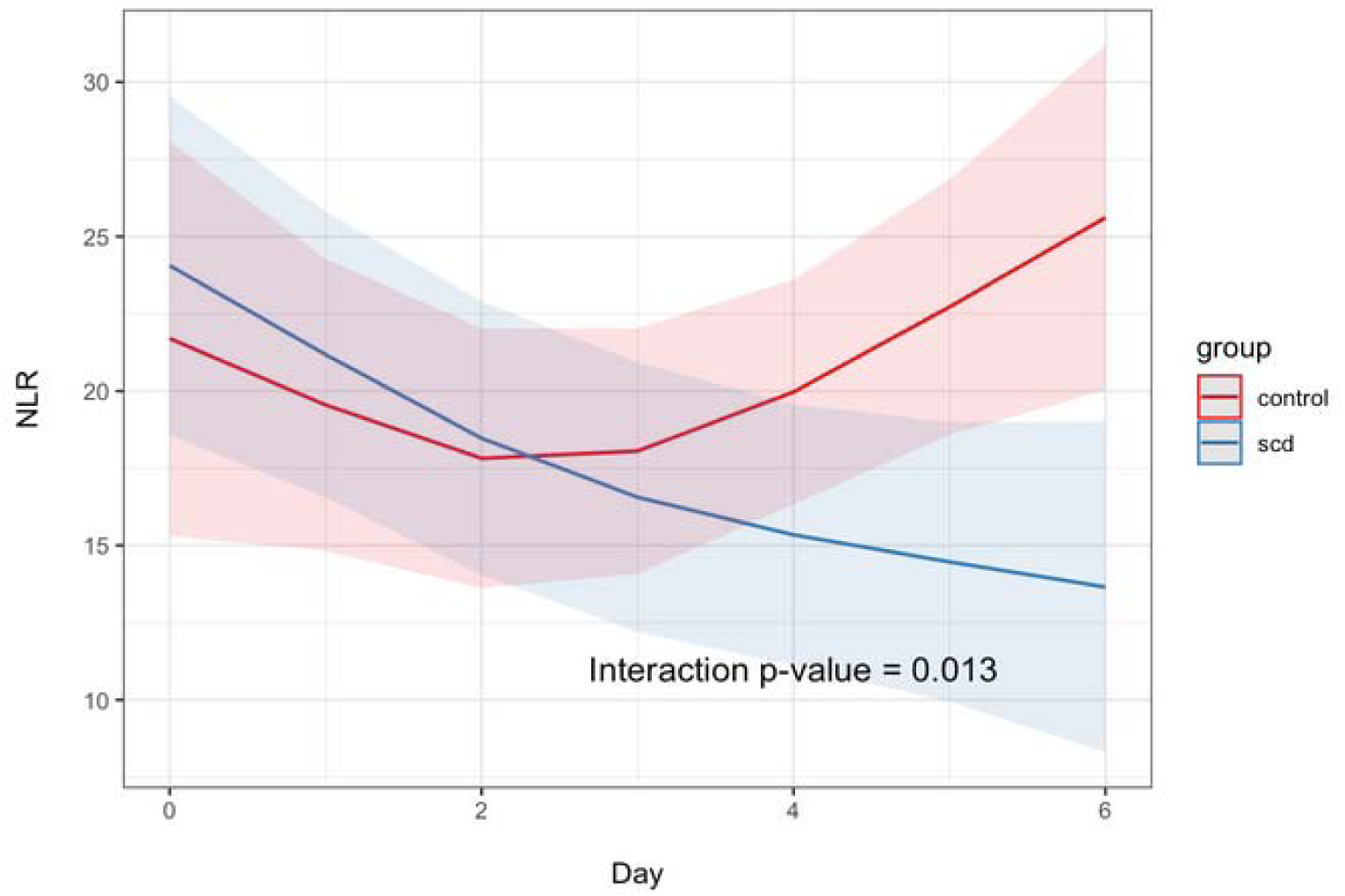
**Sensitivity analysis of SCD treatment on NLR in pooled SCD vs. control patients across adult AKI studies (China Pilot Study exclusion).**

**Figure 4:**
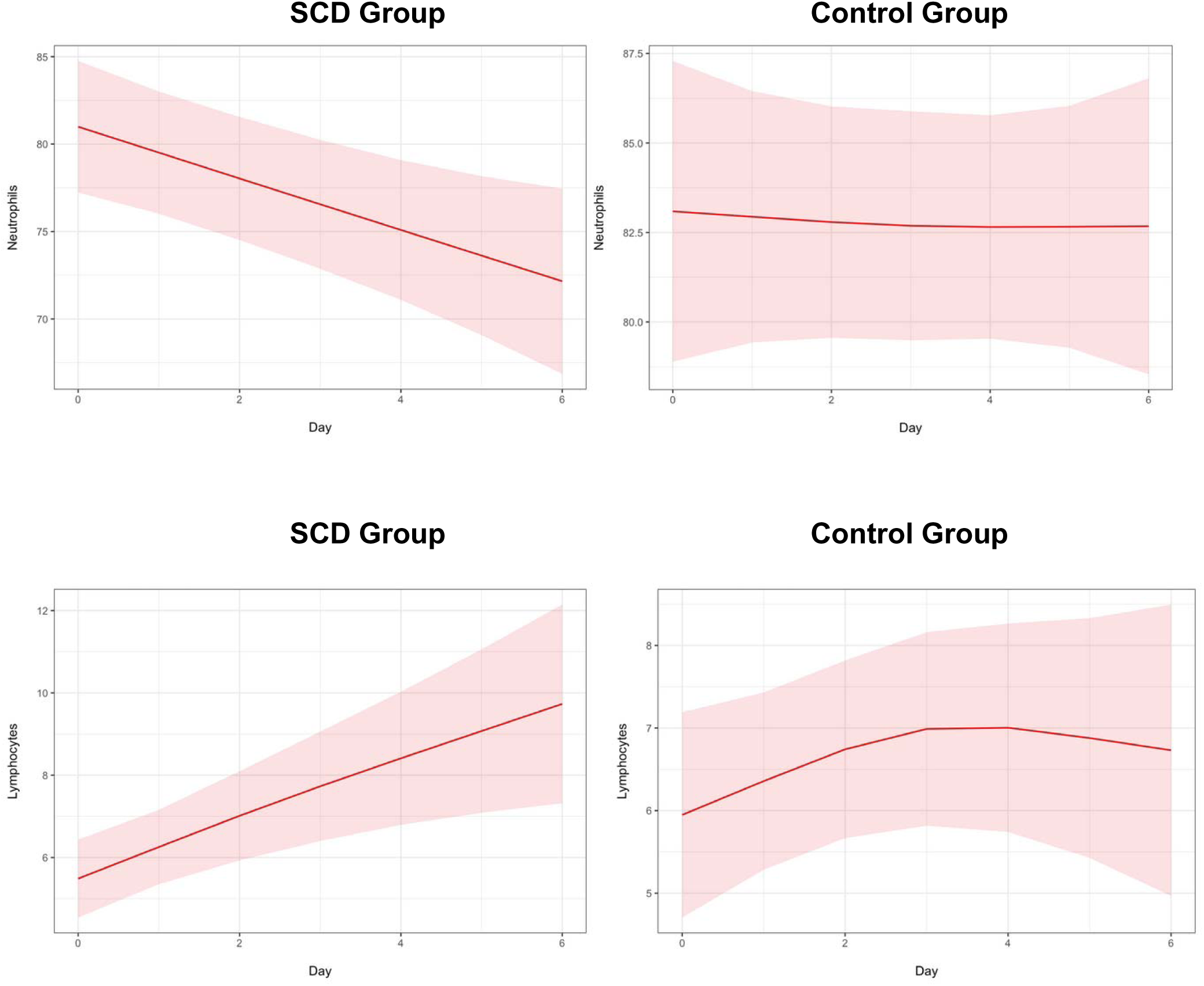
**Individual Trajectories of ANC and ALC in the Pooled Adult AKI Patient Populations.**

We examined the relationships between monocytes and lymphocytes through MLR in the pooled adult cohort. **Figure 5** shows that SCD treatment resulted in a generally static to mildly upward trend in MLR in SCD treated patients, with a more pronounced upward trend in control patients, resulting in lack of a significant difference between the 2 groups (p=0.34). Similarly, comparable curve trends and nonsignificant differences were observed in platelet counts between pooled adult SCD and control groups (p=0.12) (**Figure 6**).

## Discussion

Insights into the mechanism of the immunomodulatory effect of the SCD in dysregulated inflammatory clinical situations involving multi-organ failure, such as AKI with septic shock^29–31,38^, COVID-19^7,39^ and hemophagocytic lymphohistiocytosis^40^, have recently begun to emerge. Recently, Westover et al. reported on a series of studies that examined the mechanism of action of the SCD through in vitro bench-top assays using a mini-SCD^6^. Our findings in this analysis support the in vitro mechanistic observations by incorporating clinical pharmacodynamic data from prior SCD AKI clinical studies, further characterizing the SCD mechanism of action. The patients in these SCD AKI studies were critically ill ICU patients with multi-organ failure, the majority of whom were septic and on MV. The highly elevated baseline NLRs in adults align with previously reported ranges for AKI on CKRT (mean NLR of 21.9)^26^ and indicate elevated levels of inflammation. These values, ranging from 17 to 23, indicate significant inflammation and critical stress^41^, consistent with the highly critically ill and predominantly septic patient population in our studies (**Supplementary Table 2 and Table 1**). Similarly, the baseline NLR obtained for the pediatric cohort was also consistent with age-specific pediatric reference intervals reported for this measure^42^. Our analysis demonstrated that the SCD treatment is associated with a reduction in the NLR over time in these highly inflamed patients with AKI in both adult and pediatric trials. This reduction was driven by both a decrease in ANC and an increase in ALC in both adult (**Figure 4**) and pediatric patients (data not shown). An initial reduction in NLR was also observed in the control group, likely due to the clearance of low molecular weight inflammatory mediators via CKRT; however, this effect was not sustained, resulting in a reversal to an upward trend. The reduction in ANC aligns well with in vitro studies showing a shift in neutrophils to apoptotic senescence following SCD treatment, likely leading to a diminished neutrophil population in the body. This reduction in ANC is highly relevant when considering the critical severity and septic status (∼73-75%) of the patients with AKI in the SCD trials (**Supplementary Table 2** and **Table 1**). It is well established that delayed neutrophil apoptosis results in neutrophilia during the hyperinflammatory or “cytokine storm” phase of sepsis^43–45^. Therefore, the reprogramming of activated neutrophils to apoptosis upon SCD treatment is expected to drive a shift towards resolution of inflammation and restoration of immune homeostasis. The observed reduction of ANC in our analysis supports this hypothesis. Additionally, lymphocytes are thought to undergo significant apoptosis during sepsis, with studies showing that persistent lymphopenia significantly correlates with secondary infections and mortality ^45,46^. A recent report further demonstrated that elevated NLR was associated with lymphopenia in patients with sepsis^47^. While the exact mechanisms of hyperinflammation and immunosuppression in sepsis are still unclear, emerging evidence suggests that inflammatory neutrophils may exert a direct immunosuppressive effect on lymphocytes. For example, a recent report by Liu et al demonstrated that in a murine sepsis model, upregulation of programmed death-ligand 1 (PD-L1) (through Toll-like Receptor 2 or TLR2) on activated neutrophils, mediated by High mobility group box-1 protein (HMGB1) and other inflammatory mediators released by dead cells during sepsis, binds to programmed death protein 1 (PD-1) on T lymphocytes, leading to increased lymphocyte apoptosis^48^.

**Figure 5:**
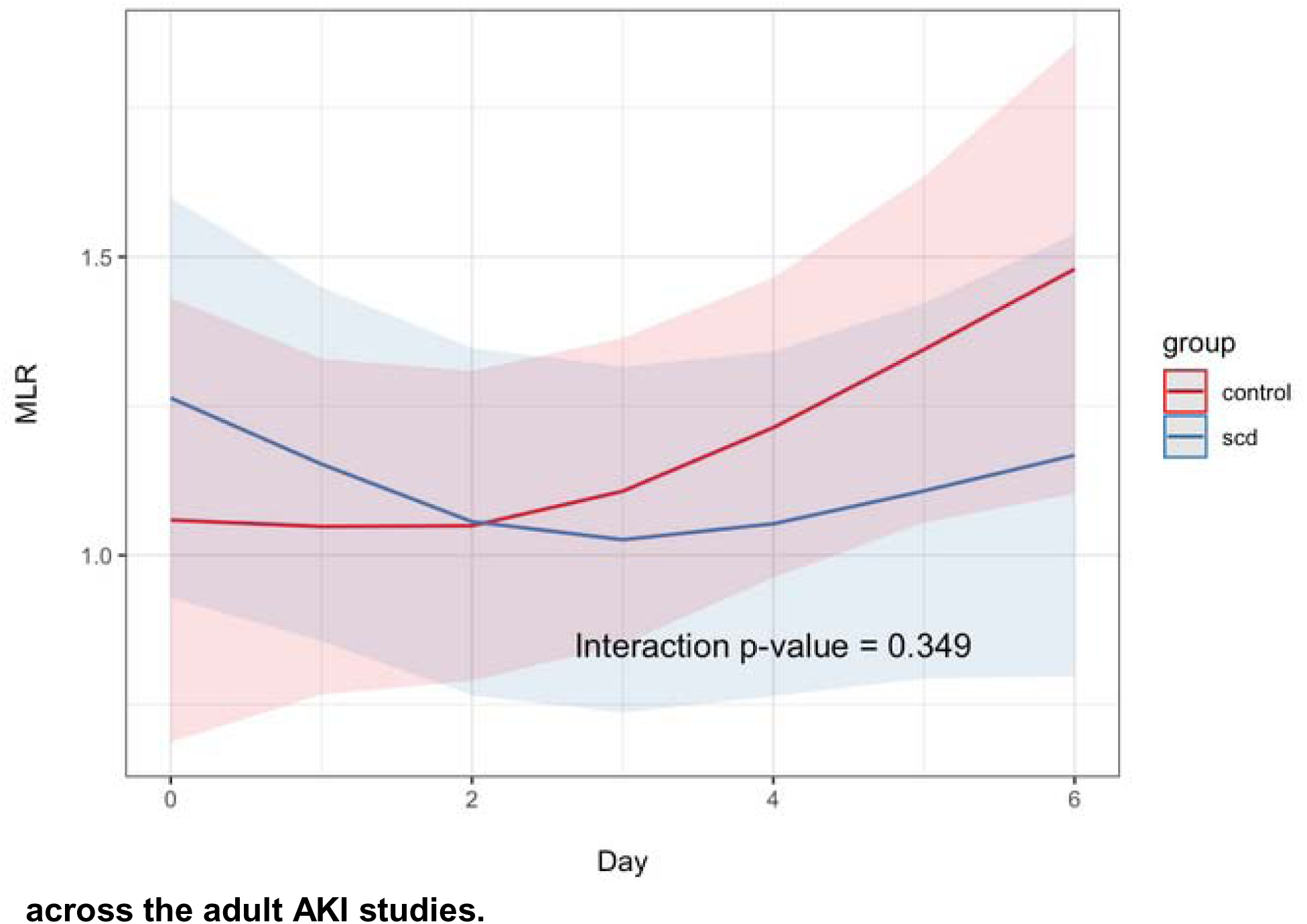
**Effect of SCD treatment on MLR in pooled SCD vs. control patients across the adult AKI studies.**

**Figure 6:**
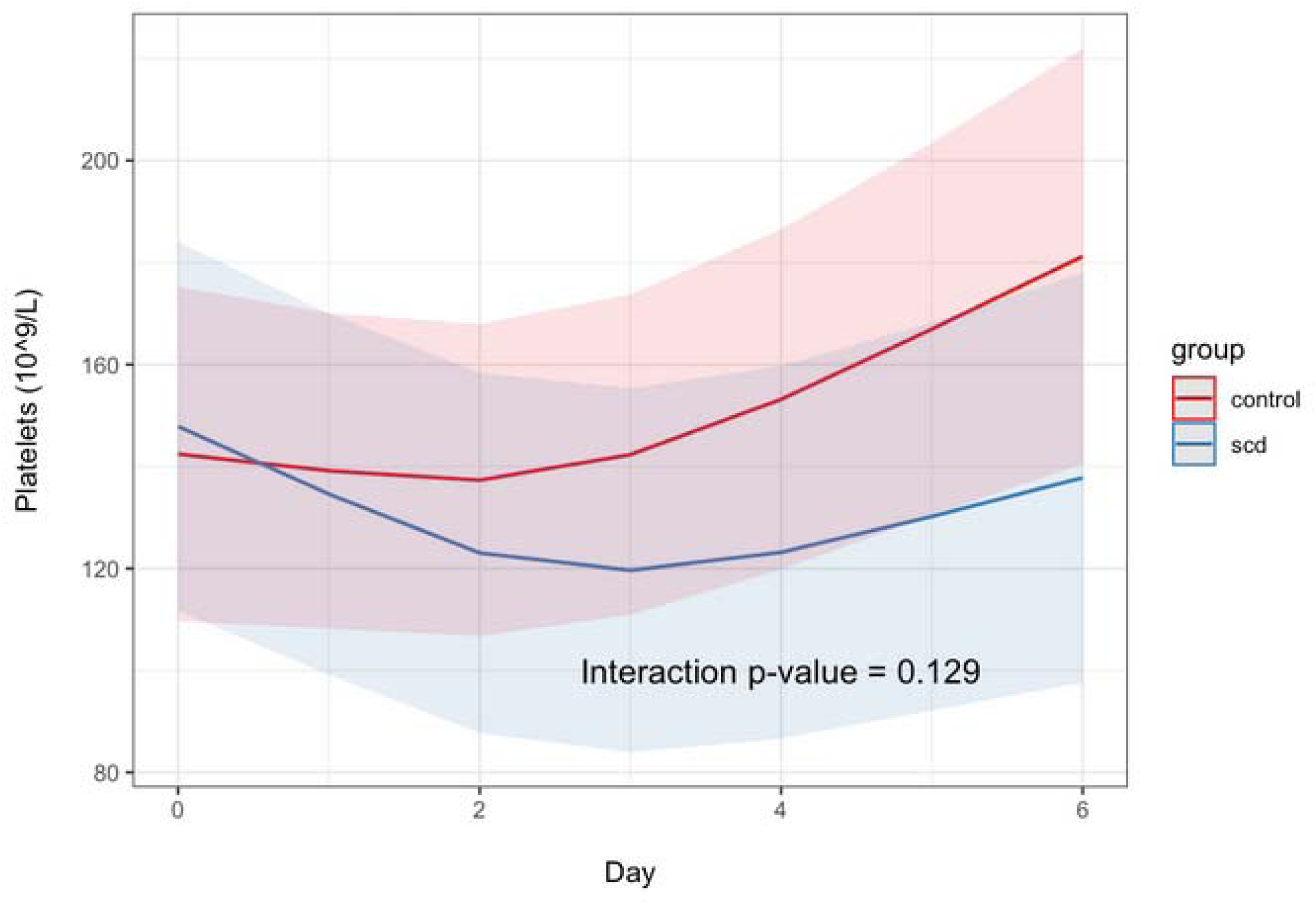
**Effect of SCD treatment on platelet levels in pooled SCD vs. control patients across the adult AKI studies.**

Regardless of the exact mechanism, it stands to reason that decreases in ANC, coupled with corresponding increases in ALC by the SCD, would signify an alleviation of the immunosuppressive environment created in sepsis, and thereby increase the likelihood of positive clinical outcomes. The increases in ALC are consistent with the lack of risk of secondary infections or device-related infections observed thus far across the SCD AKI studies^8^. Indeed, the reduction in NLR may be correlated with the positive clinical outcomes observed in the SCD AKI studies thus far, with consistent effects apparent in both pediatric and adult patient populations (**Supplementary Table 1, Figure 2**). The relative reduction of ∼44% in NLR from baseline to day 6 in the SCD-treated adult patients (**Figure 3A and B**) is consistent with the outcomes reported by Ko in CKRT survivors with AKI a which showed ∼20% reduction from peak at Day 3 to Day 5)^26^. However, NLR trajectories differed somewhat from those reported by Ko et al, where NLRs initially increased upon CKRT initiation to Day 3, with a reversal in surviving patients on CKRT back to near baseline by Day 5^26^. In our study, NLRs declined initially for both CKRT control and SCD-treated patients, with a reversal upward for control patients and a continued decline for SCD treated patients at Day 2 (**Figure 3A and B**). Reasons for this discrepancy could be differences in patient demographics, including ethnicity, and other clinical characteristics such as the timing of CKRT, the mode of anticoagulation used, and septic status. Our cohorts had nearly twice the proportion of septic patients (∼73-75%) compared to the Ko study (38.8%)^26^.

Ko and colleagues also demonstrated a linear relationship between NLR and 30-day mortality outcomes, with higher quartiles correlating with increased mortality, and greater survival in patients who experienced larger decreases in their NLR throughout their CKRT^26^. We considered a similar approach; however, we were unable to do so appropriately due to the different endpoints across the studies (discharge to ICU, 60-day mortality, dialysis dependence). Interestingly, despite the small sample size and the lack of a comparator group, it is worth mentioning that 6 of the 7 the patients who survived to ICU discharge in the China Pilot study demonstrated significant declines in their NLR from baseline, while those who did not survive experienced increases in NLR (data not shown). The fact that the effect was not observed with MLR further validated our approach and conferred further support to the in vitro bench-top MOA studies, which found that, unlike neutrophils, immunomodulation of monocytes by the SCD was through a downshift in their inflammatory profile rather than by apoptosis^6^. Therefore, the lack of a decline in MLR was not unexpected and is consistent with the findings from the in vitro studies. Similarly, the absence of differences in effect between SCD and control groups with respect to platelet levels added additional mechanistic specificity to the effect of the SCD on NLR.

Finally, this analysis is not without limitations. As mentioned above, the most significant limitation is the lack of adequate control patient data for each study. Consequently, the SCD-003 RCT served as the single major source of CKRT-only control patients. The SCD-005 control group also included CKRT-only patients from the CRRTnet registry; while this provided a valid matched cohort, it was nonetheless an external control arm. However, our pooled adult patient demographics comparison showed no differences in baseline characteristics between SCD and the control groups (**Table 3**), indicating that the clinical nature of the patients was generally similar and thus appropriate for comparison. The findings remained robust in sensitivity analyses that excluded specific cohorts. Other limitations include missing data for some patients in the studies and the lack of individual patient covariate data, as well as lack of longer term follow-up data which limited us to analysis over the 1^st^ 6 days of treatment. Since exact details of the patients whose CBC diff data were finally included in the analysis were also missing, we had to extrapolate that the clinical characteristics of the subset of patients in the final NLR analysis were reasonably represented by their larger total study cohort. However, our baseline NLR ranges and trends over time were consistent with previously reported values in patients with AKI and CKRT^26^, lending validation to our findings despite these limitations. Additionally, the lack of additional biomarker data such as inflammatory mediators, including soluble and cell surface markers of inflammation, also curtailed further characterization of the mechanistic effect of the SCD. We aim to address many of these limitations and evaluate the prognostic ability of NLR on clinical outcomes, including mortality, using data from the ongoing pivotal NEUTRALIZE-AKI randomized controlled study. This study is expected to complete enrollment in 2025, with results anticipated in 2026^49^.

In conclusion, treatment with the SCD was associated with a reduction in NLR in patients with AKI requiring CKRT and multi-organ failure, most of whom were septic. NLR is a simple, low-cost, yet highly insightful indicator of the inflammatory status that encompasses both innate and adaptive immune cell components, with significant prognostic ability demonstrated across many disease conditions, including sepsis, AKI, cancer, IBD, and neurodegenerative diseases. This analysis further supports the mechanism of action of the SCD through clinical evidence from prior AKI and sepsis studies, highlighting its potential applicability to other diseases.

## Data Availability

All data produced in the present study are available upon reasonable request to the authors

## Acknowledgments

This data was previously presented at the American Society of Nephrology’s Kidney Week 2024 Annual Meeting as abstract # 4120414 and poster # TH-PO069.

## Author Contributions

SPI contributed to the study design, hematological analysis, and drafting of the manuscript. NJO conducted the statistical analysis and contributed to the drafting of the manuscript. JLK, KKC, HDH, and LTY contributed to the critical review of the data and editing of the manuscript.

## Data Sharing Statement*

All the data in this manuscript is proprietary and owned by SeaStar Medical, Inc. All data are included in the manuscript within the figures and tables.

