## Supplemental Tables 1 & 2 for "The Impact of the Selective Cytopheretic Device on Neutrophil-to-Lymphocyte Ratios and Hematological Parameters in AKI: A Pooled Analysis"

**Supplementary Table 1: Description of Adult and Pediatric AKI studies**

| **Study Descriptor** | **Patient Population and study type** | **Study**  **N** | **CBC Diff**  **N from each study** | **Outcome Measures** | **Primary**  **Outcome** |
| --- | --- | --- | --- | --- | --- |
| **China Pilot Study^1^** | Adult AKI; single arm | 9 | 9 | In-hospital all cause mortality | 22% in SCD vs. 77.8% in matched control from PICARD registry (p=0.027) |
| **ARF-002^2^** | Adult AKI; single arm | 35 | 35 | Day 60 mortality | 31.4% in SCD vs. 50% in historical controls |
| **SCD-003^3^** | Adult AKI; randomized control trial | N=134; 69 in control CKRT only group and 65 in SCD + CKRT group | 21 from SCD riCa and 28 from control riCa groups | Day 60 mortality | 39.1% in SCD group vs. 35.6% in control group (p=0.23) (ITT analysis); 15.8% in SCD vs. 40.7% in control group (p=0.11) (PP) |
|  |  |  |  | Mortality or Dialysis Dependency at Day 60 | 16% in SCD riCa group vs. 58% in control riCa patients (p<0.01) (PP) |
| **SCD-PED-01^4,5^**  **SCD-PED-02^4,6^** | Pediatric AKI; single arm | 22 pooled patients | 22 | Day 60 mortality | 77% in SCD group vs. 51.4% in ppCRRT matched control |
|  |  |  |  | Survival to ICU discharge or day 60 | 77.3% in SCD group vs. 54.8% in ppCRRT control group (p=0.04) |
| **SCD-005^7^** | Adult COVID-19 AKI/ARDS; single arm with matched CRRTnet control cohort | N=22 all SCD group; 16 in SCD.96 and 16 in CRRTnet matched cohort | 11 in SCD group and 4 from CRRTnet matched cohort | Day 60 mortality | 50% in SCD group vs. 81% in matched CRRTnet control group (p=0.102); 31% in SCD patients treated for at least 96 hours (SCD.96) (p=0.012) |
| **Total N** |  | SCD: 153 total; 131 excluding PED studies; Control: 89 | SCD: 98 total; 76 excluding PED studies  Control: 32 |  |  |
| PP, per protocol analysis; riCa, recommended ionized Calcium range of <0.4 mM; ARDS, acute respiratory distress syndrome. | | | | | |

**Supplementary Table 2: Individual Study Baseline Mean Patient Demographics**

|  | | **ARF-002^2^** | **Pooled PED-01,02*^4-6^** | **SCD-003^3^** | | **SCD-005^7^** | | **China Pilot**  **Study^1^** |
| --- | --- | --- | --- | --- | --- | --- | --- | --- |
|  | |  |  | **SCD** | **Control** | **SCD** | **Control^‡^** | **SCD** |
| **# of Patients per Study** | | 35 | 22 | 65 | 69 | 22 | 16 | 9 |
| **Age (years)** | | 56.3 (15.0) | 9.5 (8.5) | 57.2 (13.1) | 53.5 (14.7) | 53 (17.7) | 56 (13.4) | 59.3 (13.9) |
| **Female, n (%)** | | 14 (40) | 10 (45.5) | 27 (39.1) | 25 (38.5) | 5 (22.7) | 8 (50.0) | 1 (11) |
| **Race, n (%)** | **Asian** | 0 (0) |  | 0 (0) | 0 (0) | 0 (0) | 0 (0) | 9 (100) |
|  | **Black** | 8 (22.9) | 3 (13.6) | 15 (21.7) | 14 (21.5) | 3 (13.6) | 5 (31.3) |  |
|  | **White** | 25 (71.4) | 18 (81.8) | 53 (76.8) | 48 (73.8) | 17 (77.3) | 11 (68.8) |  |
|  | **Other** | 2 (5.7) |  | 1 (1.4) | 3 (4.6) | 2 (9.1) | 0 (0) |  |
|  | **Hispanic** | 2 (5.7) | 2 (9%) | 3 (4.3) | 2 (3.1) | 1 (4.5) | 0 (0) | 0 |
| **COVID-19, n (%)** | | 0 | 0 | 0 | 0 | 22 (100) | 16 (100) | 0 |
| **Body weight (kg)** | | 95.7 (25.5) | 30.3 (33.8) | 102 (23.1) | 98.8 (23.6) | 112.4 (26.9) | 102.6 (28.5) | NA |
| **SOFA/PRISM III^†^ score** | | 11.3 (3.6) | 9.5 (7-14) | 13.8 (3.2) | 13.2 (3.7) | 11.8 (3.0) | 12.5 (2.8) | 11.0 (3.6) |
| **MV, n (%)** | | 31 (88.6) | 21 (95.5) | 61 (88.4) | 59 (90.8) | 22 (100) | 16 (100) | 4 (44) |
| **ECMO, n (%)** | | 0 | 3 (13.6) | 0 | 0 | 9 (40.1) | 4 (25.0) | 0 |
| **Vasoactive meds, n (%)** | | 31 (88.5) | 14 (63.6) | NA | | 20 (90.1) | 11 (68.8) | 6 (66%) |
| **Sepsis, n (%)** | | 28 (80) | 15 (68.2) | 45 (65.2) | 45 (69.2) | 22 (100) | 16 (100) | 3 (33%) |
| **BUN (mg/dl)** | | 46.3 (23.3) | NA | 43.2 (25.9) | 40.6 (29.7) | NA | NA | NA |
| **Creatinine (mg/dl)** | | 2.8 (1.3) | NA | 2.9 (1.7) | 2.9 (1.7) | NA | NA | NA |
| *Pooled PED cohort values with means converted from median and standard deviations from interquartile ranges / 1.35 where applicable; PRISM III at ICU Admission for PED studies; ^‡^CRRTnet matched contemporary control cohort. NA, not available; mean (SD) values represented; MV, mechanical ventilation. | | | | | | | | |
